# Signal extraction of paper-based ECG using real-world and augmented ECGs

**DOI:** 10.1101/2025.11.19.25340630

**Authors:** Nachiket Makwana, Farhan Adam Mukadam, Harshit Mishra, Pradyot Tiwari, K. Subramani, K. V. S. Hari

## Abstract

Electrocardiogram (ECG) today largely remains paper-based making them inaccessible for research and innovation. To address this unmet need, we describe an end-to-end framework for ECG signal extraction from images using a four-stage pipeline and lay down the associated data generation requirements. The signal extraction stages include perspective correction, reorientation of paper, lead isolation and signal segmentation that converts signals to voltage units enabling precise digital reconstruction of the original waveforms. Our data generation methodology generates 72,080 simulated ECG images from open source digital signal datasets. We apply novel, realistic augmentations such as coffee stains, imaging shadows and introduce 3D deformation using Blender3D. Furthermore, we employ text-to-image and image-to-image workflows by synthesizing backgrounds around ECG images using the SDXL model with IP Adapter. Our pipeline achieves acceptable signal extraction with a Pearson correlation coefficient of 0.78 and processing speeds of ≈ 1.03 seconds, which is four times faster than existing methods on a related benchmark dataset. This advancement enables practical use of ECG digitization in fast-paced clinical environments, reducing cardiology consultation turnaround times and facilitating digital storage for population health analysis.

## 1. Introduction

An ECG provides clinicians with their first glimpse into cardiac function of a patient, making it indispensable for initial cardiovascular assessment. A 12-lead ECG enables rapid treatment decisions by detecting arrhythmias, myocardial infarctions, structural heart abnormalities, electrolyte imbalances, and medication effects. It has been demonstrated that deep learning models can assess ECGs and detect critical conditions rapidly, which can improve patient outcomes. Previous studies [1] [2] [3] have demonstrated that these models can detect important cardiac conditions and some even go beyond conventional inferences such as estimating ejection fraction [4].

To facilitate ECG analysis using deep learning in clinical practice, a digital signal is required which is often unavailable due to hardware limitations. Most of the healthcare facilities still rely on older ECG machines that lack digital export capabilities or store outputs as rasterized PDFs. This highlights a fundamental gap between the digital requirements for automated analysis and the analog reality of current ECG documentation practices. To enable widespread adoption of AI-based cardiac assessment tools, bridging this format incompatibility is essential.

Several approaches have already been developed to address ECG signal extraction, ranging from synthetic data generation to fully automated pipelines. However, most of these methods require human intervention at some stage which makes them difficult to deploy. Only recently some deep learning based solutions have demonstrated pipelines which do not require any human intervention making them a good choice for clinical settings, however they need to be evaluated thoroughly for clinical settings.

### Simulated data generation

Basic image generation of signals can be easily done using plotting libraries [5] like ecg-plot by Dy [6] and ecg-image-kit by Shivashankara et al. [7]. Both libraries convert raw 12-lead ECG signals into simulated images. The ecg-image-kit library applies augmentations to mimic real world imaging scenarios but still incorporates only 2D transformations which may not be enough to encompass real world scenarios of 3D deformations. Demolder et al. [8] has used a similar simulation-based generation approach using about 100 samples from PTB-XL dataset [9] to evaluate their digitization pipeline. Sangha et al. [10] employed ecg-plot to create images from CODE [11] and PTB-XL datasets and trained CNN models to classify these ECGs across six conditions using similar augmentation techniques. Almost all of these approaches do not consider the background conditions of the ECG paper which is assumed to be clean but in critical care settings there can be lot of clutter making the paper localization itself a tough task.

### Document rectification

Mobile imaging of ECG thermal papers introduces additional challenges, particularly paper warping from storage in files or rolls, which creates distortions during analysis. While not being ECG-specific, document rectification research has developed relevant approaches for handling such deformations. Das et al. [12] created the Doc3D dataset to train a dewarping network which addresses paper folds, creases, and surface bends. Similarly, Hertleim et al. [13] proposed a Blender-based pipeline for generating coupled deformed images and correction data for training dewarping neural networks. These document rectification techniques provide a foundation for addressing similar challenges in ECG image processing.

A critical challenge in developing ECG signal extraction systems is the scarcity of paired datasets containing both ECG images and their corresponding digital signals, leading researchers to rely heavily on simulated data generation approaches.

### Real-world paired ECG data

While Randazzo et al. [14] and Fortune et al. [15] obtained real-world ECG image-signal pairs from hospitals, these datasets lack essential metadata such as lead bounding boxes or signal masks. Although they remain valuable for evaluation purposes, the absence of annotations limits their utility for training digitization models.

### ECG signal extraction

ECG signal extraction research has received less attention than disease detection from digital ECGs. Most approaches adopt multi-stage pipelines involving skew correction, lead isolation, binarization, and voltage scale conversion. However, many methods require manual interventions such as lead cropping or threshold selection [14] [16] [17], limiting their scalability in clinical settings. Another problem with such approaches is that they conform to specific lead layouts and are not robust against layout changes.

To overcome these challenges with basic image processing approaches, deep learning has been incorporated into digitization pipelines which are semi-automated in nature. Mishra et. al. [18] used deep learning to predict the correct binarization threshold for signal segmentation. Another approach by Li et. al. [19] treated digitization as a semantic segmentation problem, and proposed this as an end-to-end deep learning approach for signal extraction from noisy paper records. Shivashankara et. al. [7] train a Denoising CNN for grid removal, a YOLOv7 model to detect regions of interest (rows of ECG leads) to isolate the leads and then scanning the lead image left to right to extract the signal. These approaches, while being viable do not mention about intersecting signals which is encountered often in clinical settings.

The most recent fully automated approach proposed by Demolder et. al. [8] employs a two-stage deep learning pipeline for end-to-end ECG digitization. The first stage involves identifying grid intersections, correcting deformations, and calibrating axes. Then the second stage detects waveforms as masks and applies post-processing to generate digital signals. The workflow is sound but the validation is conducted on a curated dataset and real world imaging could be more messy.

### Our contribution

We demonstrate a complete framework for ECG signal extraction that drives each part of our dataset generation process, where signal extraction stages motivate the corresponding data generation needs. The signal extraction process requires: (1) segmentation of the region of interest (i.e., the ECG paper) to eliminate perspective issues, (2) isolation of leads using oriented bounding boxes, and (3) segmentation of each signal to convert back to true voltage values. To address these requirements, we generate 72,080 images - 62,700 base images with the corresponding lead bounding box and signal masks - and the remainder as 3D deformations without coupled ground truths. Given diverse backgrounds in real-world imaging, our generation approach employs 3D software and novel synthetic background generation techniques to facilitate the ECG paper segmentation step. A detailed description of the signal extraction methodology and its corresponding data generation process is provided in the methodology section.

## 2. Methodology

Our methodology consists of two components: simulated ECG data generation and the signal extraction pipeline described in their respective sections.

### Section I - Dataset generation

The aim of the data generation process is to mimic real-world mobile imaging conditions including warping, lighting variations, paper mutilations, camera angles, and paper deformations. We employ PTB-XL and MIMIC-IV ECG [20] datasets with 20,000 signals chosen randomly from both datasets combined to generate images using ecg-image-kit and a modified ecg-plot library. We enhanced ecg-plot to include DC pulse generation, additional grid colors, long ECG strips, and coupled signal bounding boxes with lead masks. This constitutes our base image generation. Figure 1 summarizes the complete data generation pipeline in the left side. Figure 2 shows representative outputs from both libraries with minimal augmentation.

**Figure 1.**
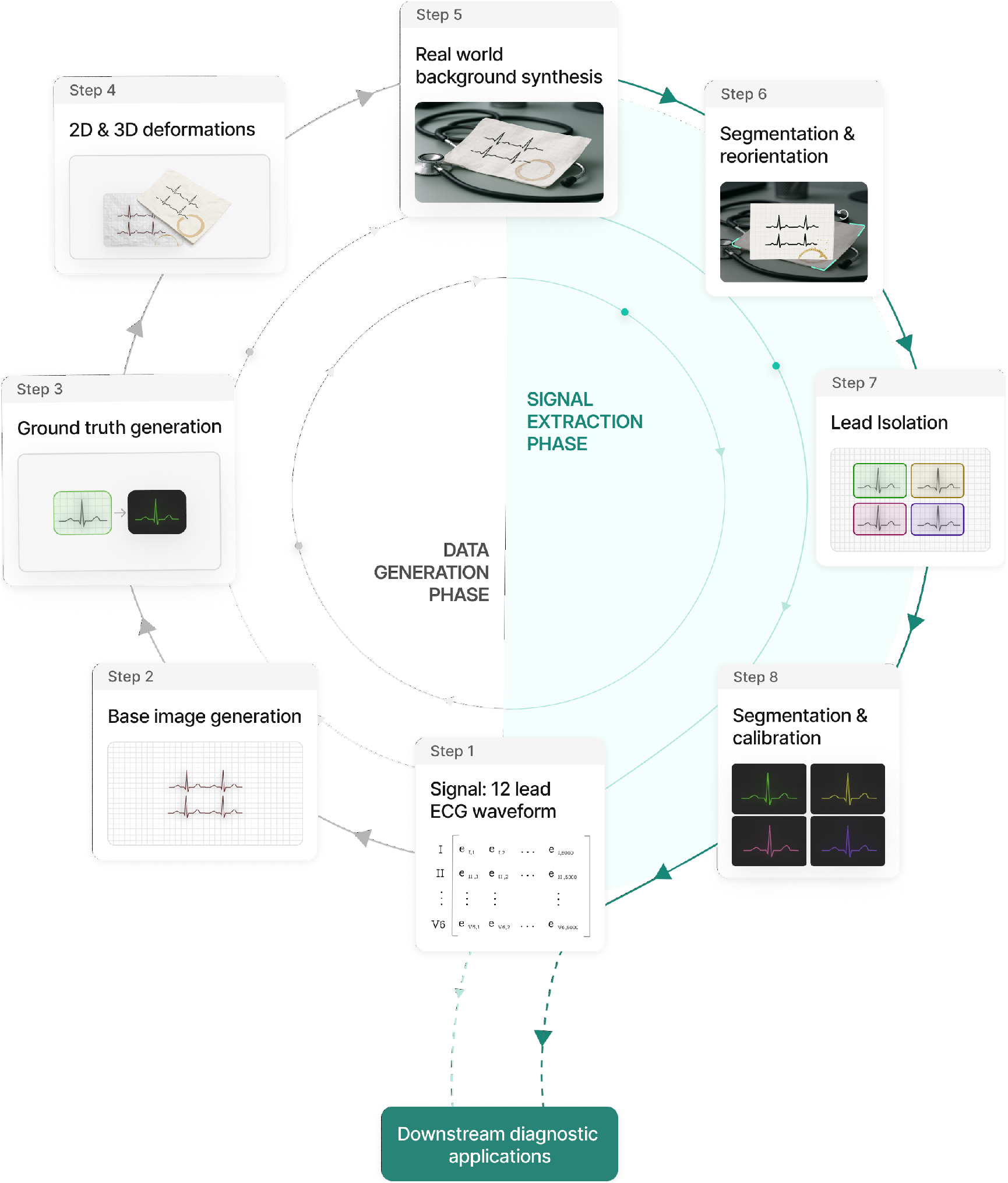
Comprehensive framework for generating synthetic ECG datasets and extracting signals for downstream diagnostic applications.

**Figure 2.**
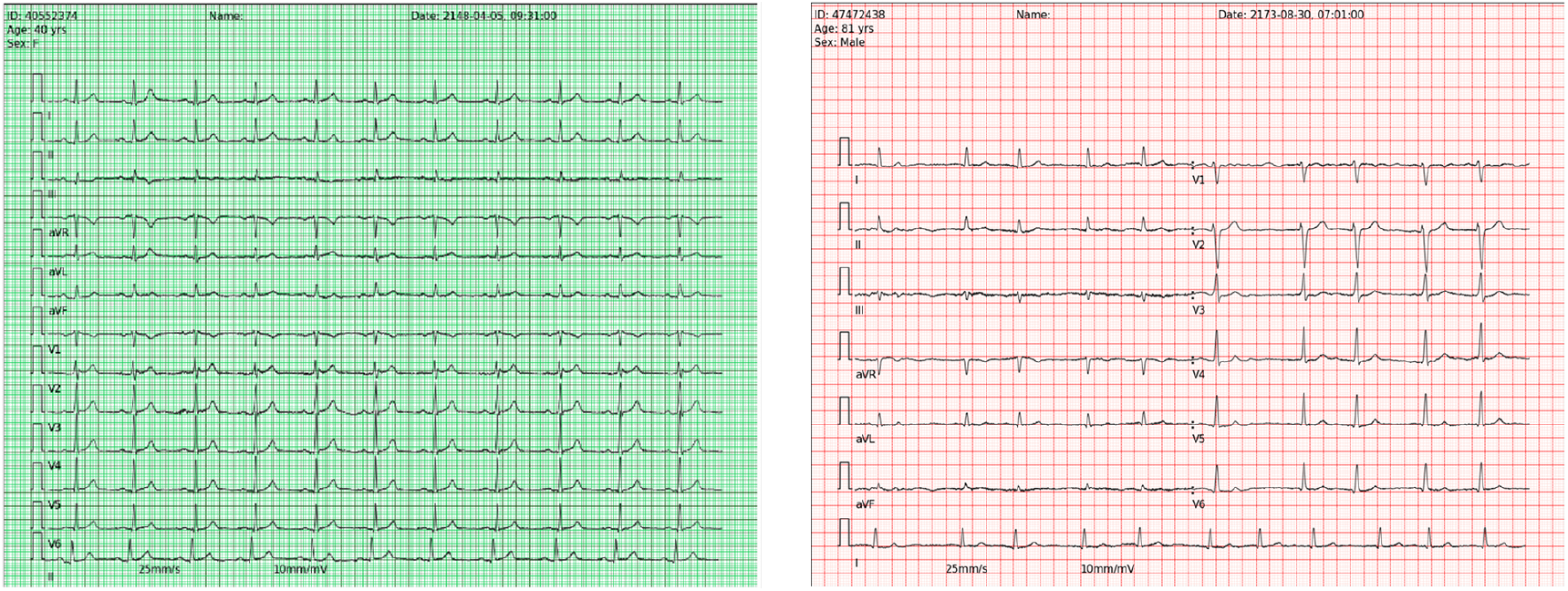
Example outputs of base image generation.

The initially generated images had the drawback of appearing too pristine and resemble scanned documents rather than real-world mobile captures. We therefore apply custom augmentations designed after analyzing real-world ECG images. Our novel augmentation pipeline includes overlaying transparent liquid and coffee stain images to simulate common paper degradation and choosing grid colours which simulate faded colour ink. Figure 3 demonstrates these augmented images with various transformations applied.

**Figure 3.**
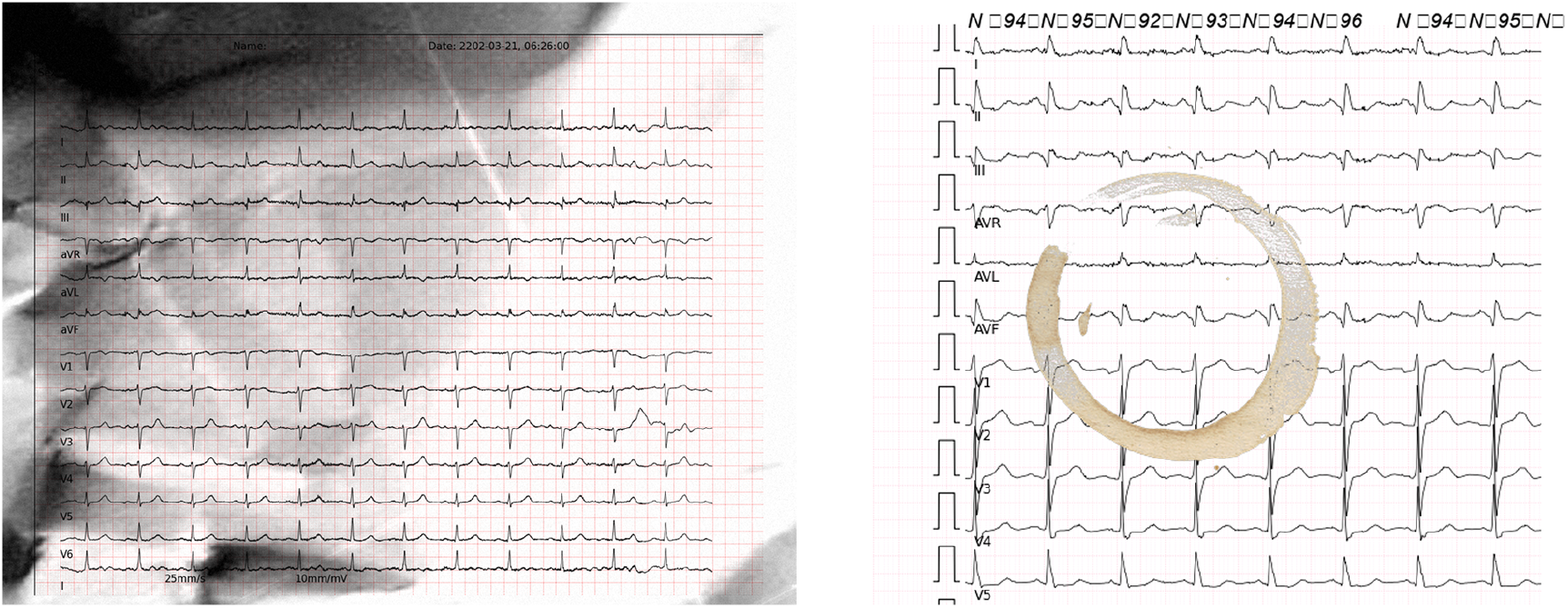
Augmented images with various transformations applied including coffee mug stain overlay.

This controlled generation approach produces coupled ground truth annotations for training multiple digitization pipeline stages. Each 12-lead ECG generates comprehensive annotation data including oriented bounding boxes and signal masks. This ground truth data is stored as JSON files for bounding boxes and PNG images for individual lead masks. Figure 4 shows a representation of the corresponding ground truth oriented bounding boxes and signal masks. Base images combined with their augmentations constitute majority of our data repository.

**Figure 4.**
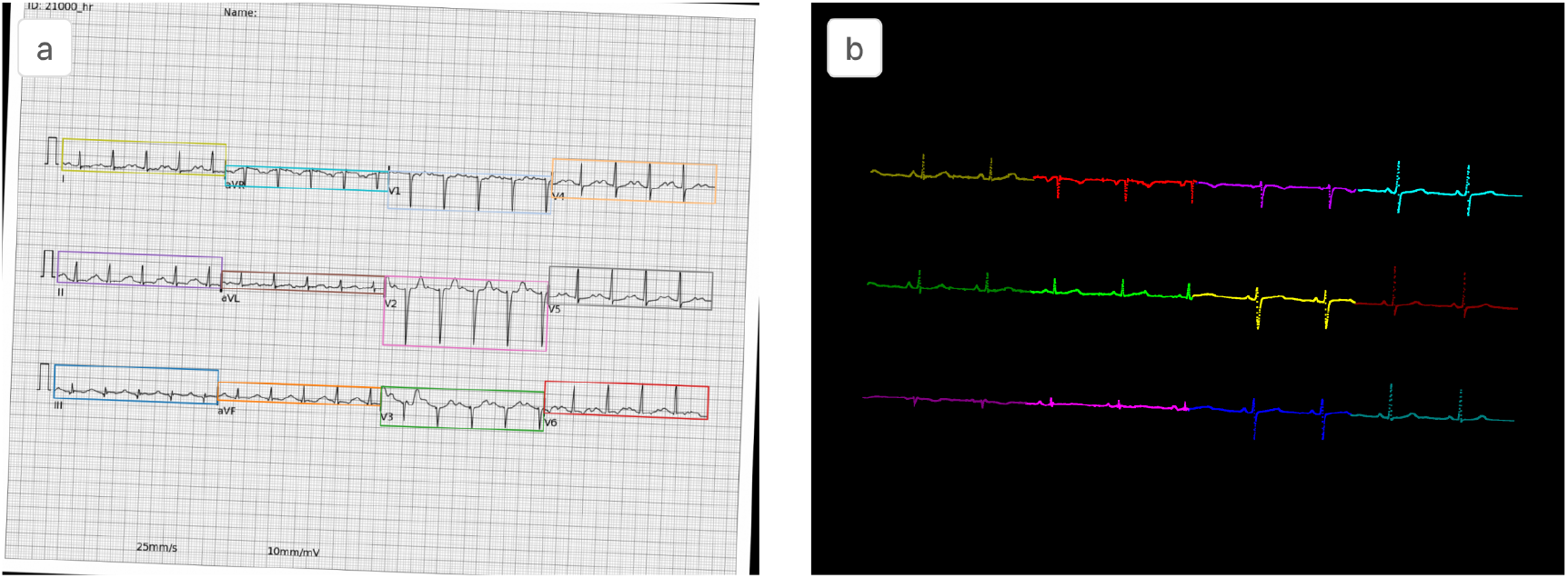
Ground truth annotations for a single 12-lead ECG. (a) ECG image with oriented bounding boxes for each lead, (b) Signal masks for all leads.

Most of our expected input is mobile imaging which introduces paper warping that causes signal distortion. We address this by applying 3D deformations using Blender3D [21] to simulate realistic paper warping conditions. ECG images are mapped as textures onto 3D planes and transformed using parametrically controlled deformations to generate realistic image variations. Figure 5a. The pipeline also simulates a commonly observed scenario of phone and hand shadows during image capture as seen in Figure 5b

**Figure 5.**
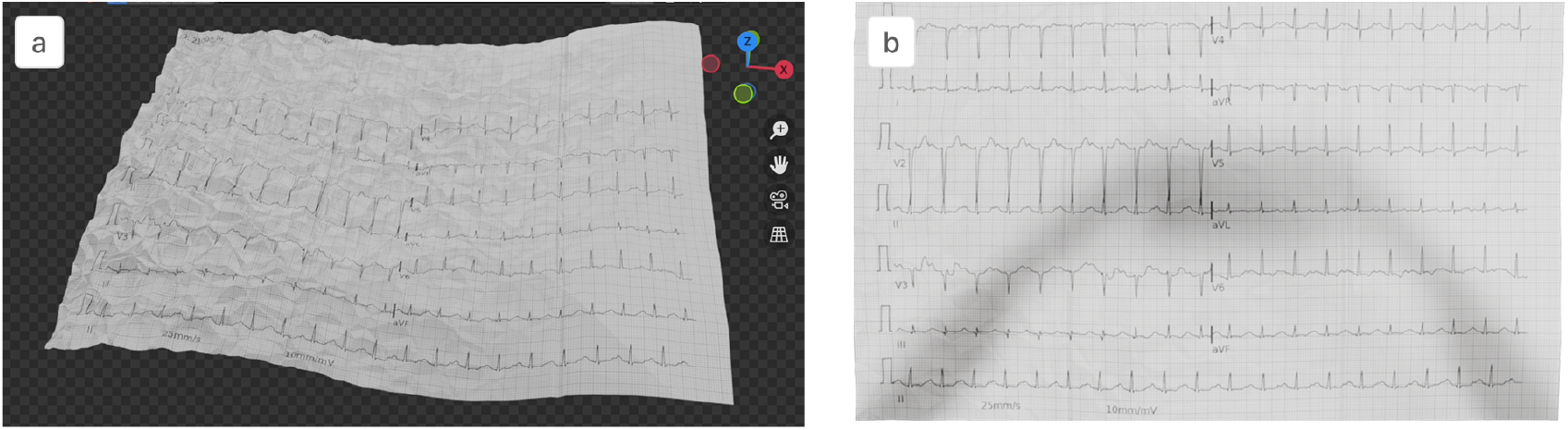
Images with 3D deformation. (a) Deformations introduced in Blender 3D. (b) Simulated phone and hand shadows.

To enhance realism further through we do background synthesis using Stable Diffusion XL (SDXL) [22] with IP-Adapter [23]. Our initial experiments showed that SDXL alone was not enough to generate realistic backgrounds with text prompts only. Hence we curated 20 reference images representing typical ECG environments including examination tables, ICU settings, wooden surfaces, and file folders. We use IP-Adapter to provide these reference images as guidance for the background generation. Each reference generates diverse backgrounds with coupled text prompts. The transparent 3D-deformed ECG images are then composited with these simulated backgrounds, creating datasets that mirror real-world mobile imaging scenarios. Figure 5 shows 3D deformation examples, while Figure 6 demonstrates the complete background synthesis pipeline.

**Figure 6.**
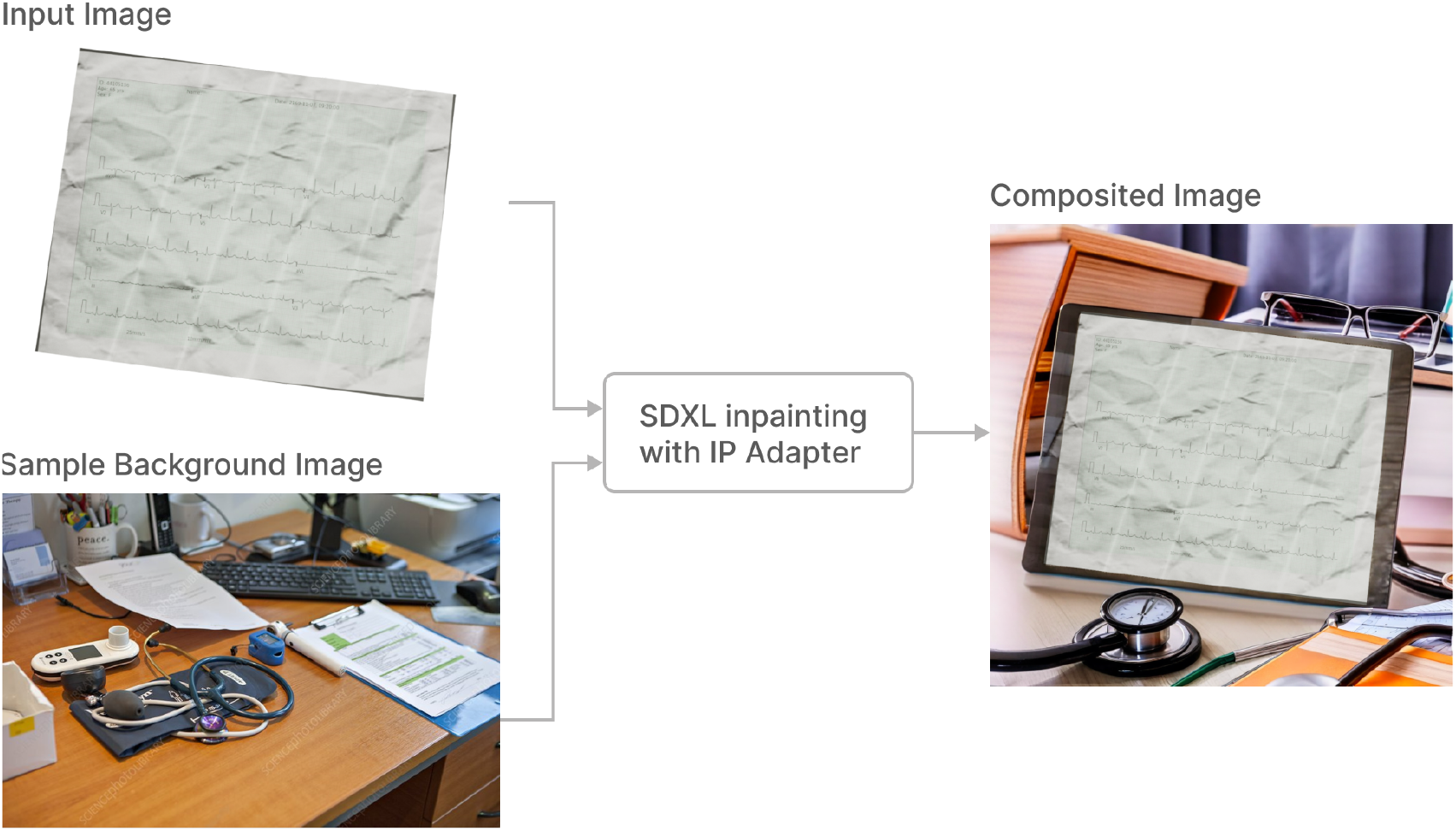
Background synthesis workflow

The total count of images with ground truths is as seen in Table 1. Using the comprehensive simulated dataset described above, we develop our four-stage digitization pipeline. This automated workflow processes real-world ECG images captured under suboptimal conditions, requiring no user intervention to enable focus on patient care rather than image quality.

**Table 1:** Generation stages and dataset statistics.

| Generation Stage | Description | Count |
| --- | --- | --- |
| ECG image generation | Standard ECG plots with signal masks and bounding boxes with augmentations included | 20000 |
| ECG image generation with augmentations | Extended ECG strips annotations and augmentations generated using Albumentations library | 42700 |
| ECG generation using 3D pipeline | 3D transformed images without backgrounds using blender with synthesized backgrounds. | 9380 |
| **Total Unique Images** | Combined dataset for model training | **72080** |

### Section II - Signal extraction pipeline

The signal extraction pipeline comprises: (1) perspective correction through paper segmentation and corner warping, (2) image orientation correction, (3) lead isolation using oriented bounding box detection and (4) signal segmentation on cropped lead patches with voltage calibration. Figure 1 illustrates the complete workflow in the right panel.

#### Segmentation of paper

We train a semantic segmentation model to identify paper boundaries for perspective correction. The model uses our 3D deformation dataset, where alpha channels are converted to binary masks paired with background-synthesized images. We adapt the FFCResNet [24] (Fast Fourier Convolution Residual Network) architecture from inpainting to segmentation tasks. Using the contour of the mask, we do a perspective transformation to correct viewing the angles in mobile images. For images which are already aligned, we include scanned/pristine imaging in the training process where the entire image area is predicted as a foreground mask and it does not require transformation. Figure 7 demonstrates perspective correction applied to real-world ECG captures.

**Figure 7.**
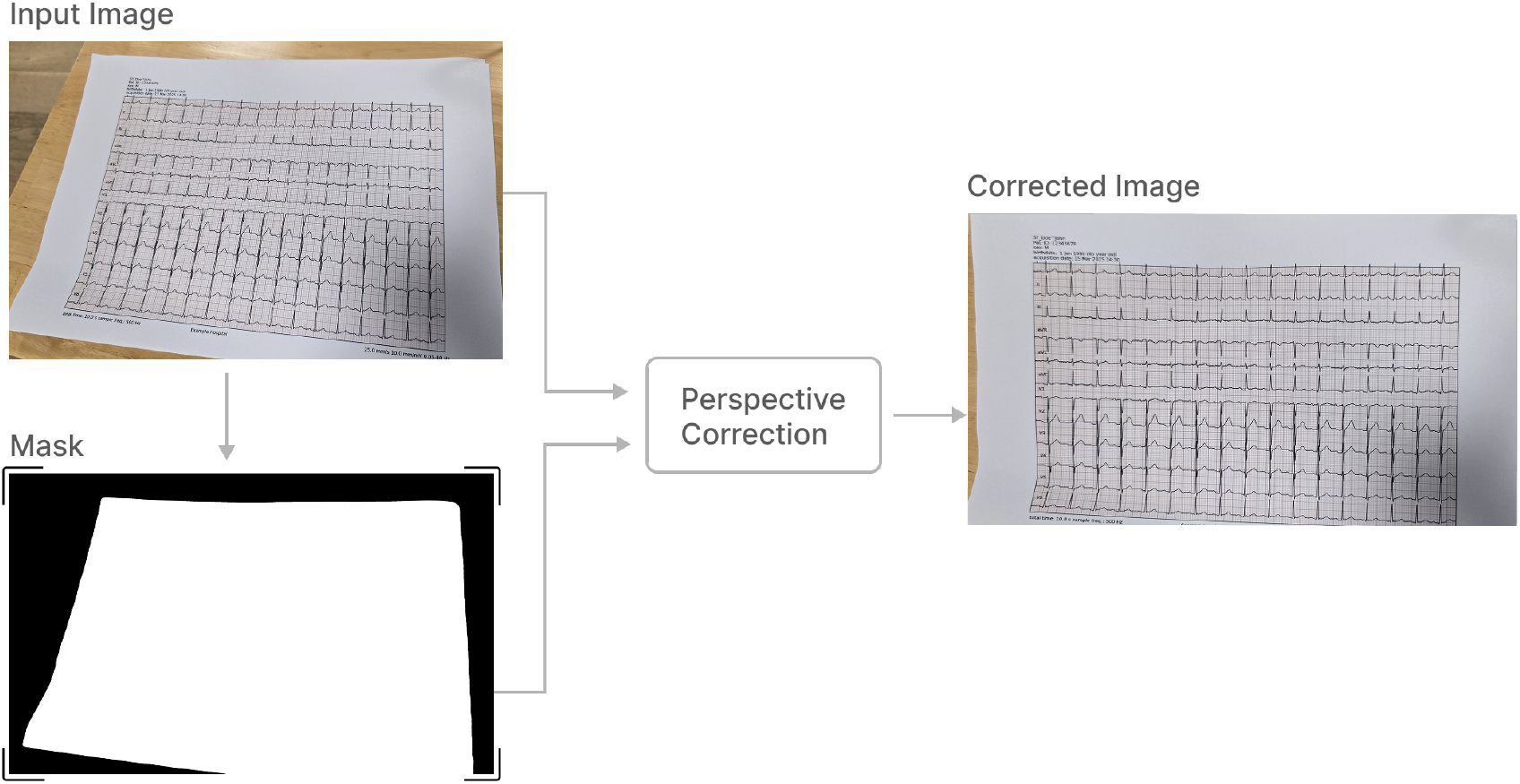
Perspective correction

#### Reorientation of paper

Following perspective correction, we address image orientation variability. Clinical settings often produce rotated images due to urgency or spatial constraints. We train a DenseNet121 [25] classifier to detect rotation angles (0°, 90°, 270°) and apply corresponding corrections. This provides coarse orientation adjustment, with fine-grained alignment handled by oriented bounding box detection (OBB) in the subsequent lead isolation stage.

#### Isolation of leads

We train a YOLOv11 OBB model [26] using our coupled dataset to detect and classify all 12 ECG leads (I, II, III, aVR, aVL, aVF, V1, V2, V3, V4, V5, V6). Initial training on 60,000 simulated images showed limited performance on real mobile captures. Adding 120 manually annotated diverse scenarios significantly improved detection accuracy. We observed minor issues with the OBB output which needed some postprocessing to ensure all bounding boxes are of the same length and aligned properly. This post-processing step applies DBScan clustering to bounding box centroids, automatically determining column layout without prior specification of cluster numbers. This column layout determination helps in the timing calibration during the final digitization step. While the dataset does contain full mode/rhythm signals, we currently do not consider them during the digitization and remove them during postprocessing even if detected. Figure 8 shows OBB predictions on real-world imaging.

**Figure 8.**
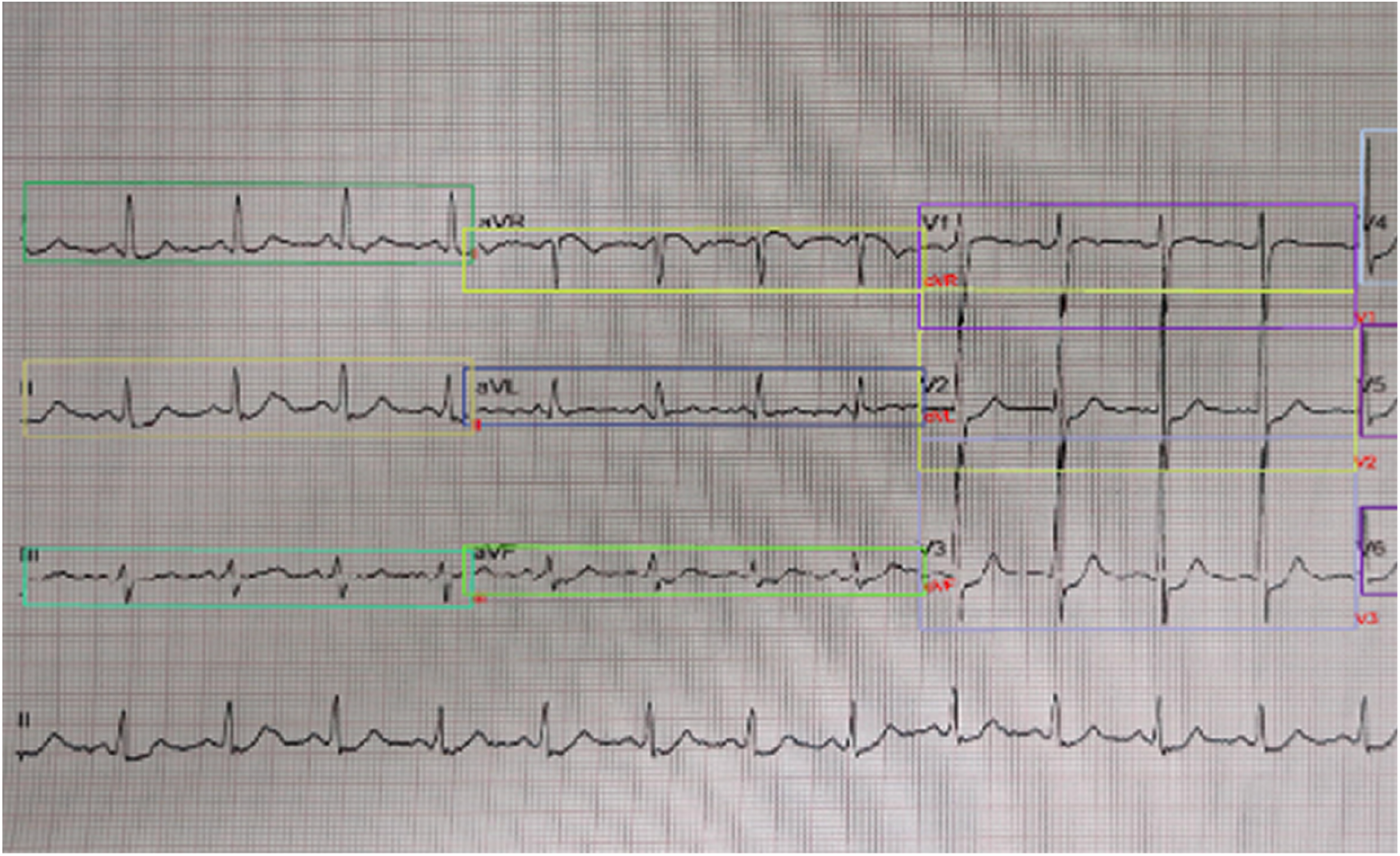
Inference on real world imaging (Lead annotations on the right of the boxes)

#### Extraction of signal

We crop individual leads using OBB coordinates and apply FFCResNet segmentation to extract signal traces. Each cropped lead is divided into 512×512 pixel patches with black padding and centered positioning. This patching strategy captures fine signal geometry while accommodating varying image resolutions. The model predicts signal masks for each patch, which are then stitched to reconstruct complete lead masks. Post-processing removes noise from neighbouring lead regions which appear in the cropped lead image. To handle signal overlap, we trained the model using our customized base image data with configurable lead spacing, enabling the model to distinguish true signals from adjacent traces despite visual overlap in source images. Figure 9 demonstrates an example of patch based segmentation results for one lead. Segmentation masks are converted to digital signals by scanning each lead image left-to-right and recording Y-coordinates of the first white pixel in each column. This produces pixel-based signal coordinates that require calibration to voltage units. We estimate signal timing using the column count from OBB detection, then calculate voltage scaling from lead image dimensions and timing parameters. All voltage calculations are done by considering paper speed 25mm/s and 10mm/mV as default parameters. An estimated central baseline enables correct positive and negative deflection mapping. The resulting 12-lead signals are exported in CSV or DICOM format for downstream analysis including fiducial point detection, interval measurements (RR, PR, QT), and clinical condition assessment.

**Figure 9.**
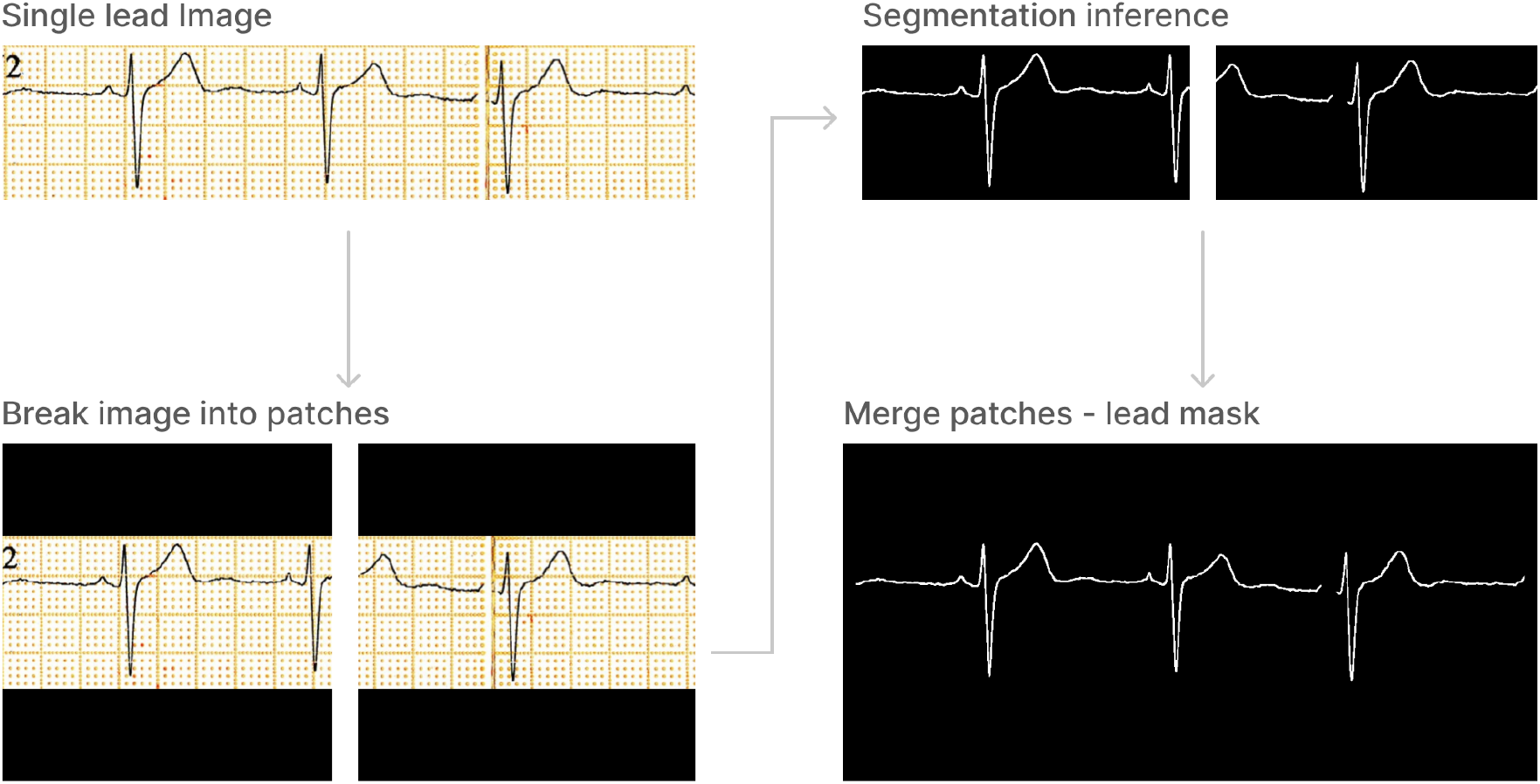
Patch based semantic segmentation

This complete pipeline enables fully automated ECG digitization from mobile images, producing digital signals without manual intervention.

## 3. Results

We evaluate digitization performance using the PMcardio ECG Image Database (PM-ECG-ID) [27], which provides deformed ECG images with corresponding ground truth signals from PTB-XL. Our evaluation focuses on 5,640 images containing complete 12-lead layouts from the total 6,600 image dataset. The evaluation revealed some temporal misalignment between predicted and ground truth signals despite signal morphology and amplitude being preserved. To mitigate this issue we apply Dynamic Time Warping (DTW) for temporal synchronization before computing metrics. An example of the ground truth and predicted signal after time alignment is shown in Figure 10 To compare with Demolder et al. [8], we report Mean Absolute Error (MAE), Root Mean Square Error (RMSE), Signal-to-Noise Ratio (SNR), and Pearson’s Correlation Coefficient (PCC) across the dataset (Table 2). SNR is calculated using the following equation.

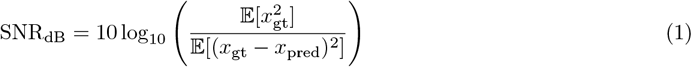

**Table 2:** Performance comparison between our method and baseline approach.

| Metric | Our Method | Demolder et al. [8] |
| --- | --- | --- |
| MAE (mV) | 0.069 | - |
| RMSE (mV) | 0.132 | 0.056 |
| SNR (dB) | 8.471 | 15.050 |
| PCC | 0.781 | 0.948 |
| Processing Time (s) | $1.03 \pm 0.03$ | 4.86 |

**Figure 10.**
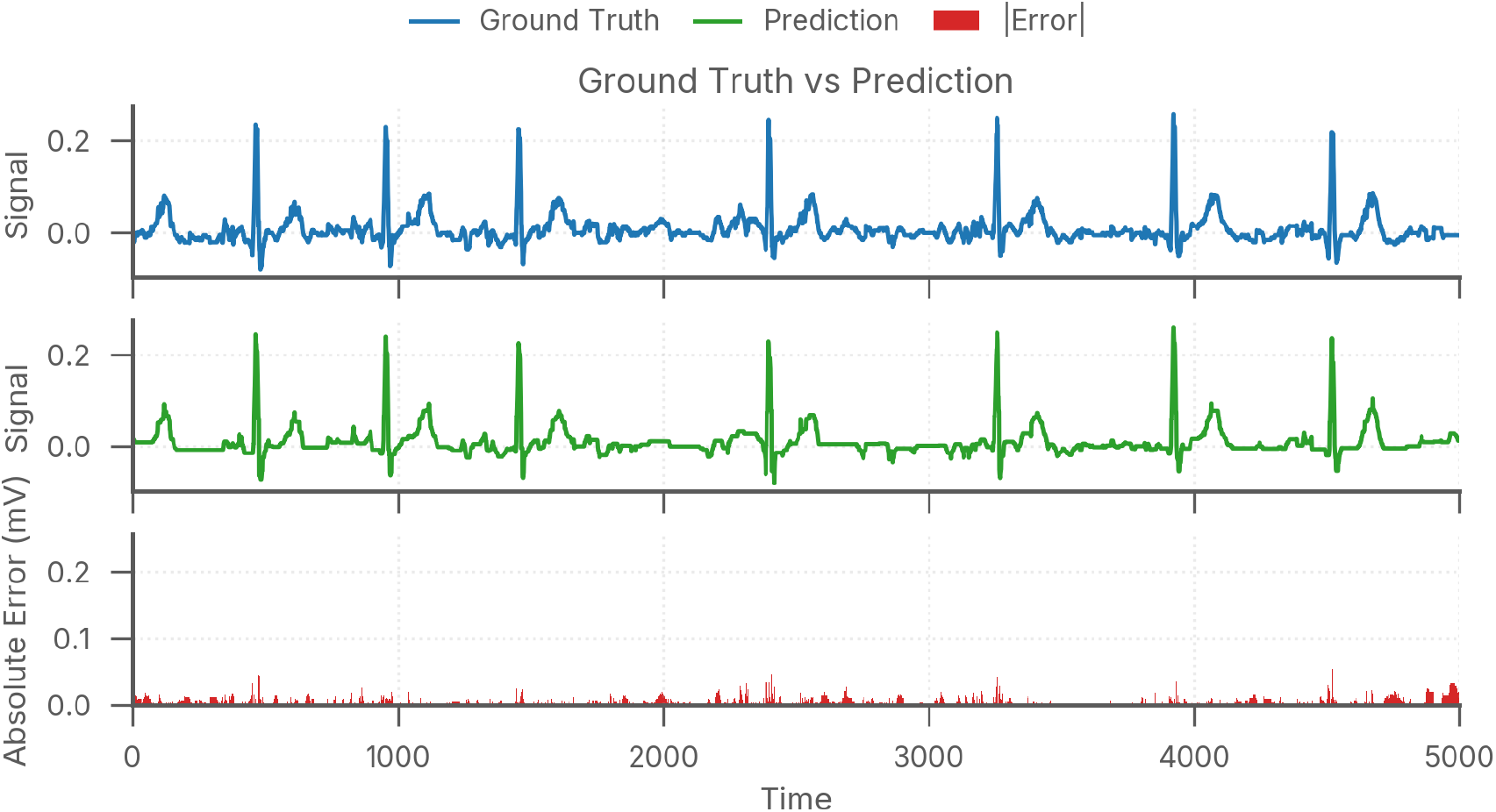
Ground truth and Predicted signal comparison. The red error bar shows the sample wise absolute difference in the Ground truth and predicted signals.

Where 𝔼 [] denotes the expected value (sample mean), *x*_gt_ is the ground truth signal, and *x*_pred_ is the predicted/reconstructed signal.

Our pipeline achieves significantly faster processing, completing the entire digitization workflow in 1.03 seconds per image compared to 4 seconds as reported by Demolder et al. [8], which demonstrates practical advantages for clinical deployment. Our pipeline targets mobile imaging scenarios with greater variability in lighting, perspective, and paper deformation, whereas prior work focuses on controlled scanning conditions. The moderate correlation (PCC = 0.748) indicates preserved signal morphology despite quantitative differences.

## 4. Discussion

We have demonstrated a working pipeline for converting EGG image back to signals. Our evaluation demonstrates the trade-offs inherent in fully automated ECG digitization for mobile imaging. While quantitative metrics show performance gaps compared to Demolder et al. [8], our approach addresses fundamentally different challenges in real-world deployment scenarios.

Synthetic data generation addresses similar challenges across domains including satellite imagery [28], civil engineering [29] [30], robotics [31] [32], and fashion [33], where coupled ground truth data is expensive or impossible to collect at scale. On similar lines, we structure our dataset generation methodology to leverage 3D modeling in conjunction with generative AI techniques to construct comprehensive training datasets for ECG digitization tasks.

Our method highlights three key innovations: Stable Diffusion based clinical background synthesis, novel augmentations including coffee stains and imaging shadows, and finally optimized processing achieving 4x speed improvement.

We do address perspective and orientation correction but do not implement explicit dewarping of paper folds. This design choice maintains processing efficiency while our 3D deformation training data provides the foundation for future dewarping integration.

Existing digitization approaches exhibit several methodological limitations. Prior studies have largely relied on controlled imaging conditions using dedicated scanners [14][16][17], which restricts their use in real clinical settings. The semi-automated approaches that demand manual lead cropping or threshold adjustments [22,24,25,26] prove unsuitable for emergency situations requiring immediate ECG interpretation. While recent deep learning methods [18][19] have advanced signal extraction capabilities, they fail to address the significant imaging variability encountered when ECGs are captured using mobile devices.

One could argue that skipping the digitization process altogether and using direct CNN-based disease classification from ECG images [10] could reduce all this overhead but it can have significant limitations. Low-resolution inputs (224-512 pixels) are standard input sizes for such models which may obscure critical features like ST-segment changes, while the black-box nature limits clinical explainability. Our digitization approach preserves signal fidelity and enables traditional ECG analysis workflows.

Future enhancements could address current limitations through several approaches. Integrating dewarping using our existing 3D geometric metadata would handle severe paper deformations such as crumbles or folds because of paper filing. Additionally, large vision-language models present an opportunity for an end-to-end signal regression experiment which could bypass segmentation and post-processing stages entirely. Such approaches could directly predict voltage arrays from lead images, simplifying the pipeline while leveraging advances in foundation model capabilities.

## 5. Conclusion

We present a fully automated ECG digitization pipeline that processes mobile-captured images without manual intervention. Our comprehensive simulation framework combines 3D deformation modeling, clinical background synthesis, and novel augmentations to train robust deep learning models. Our deep learning based digitization pipeline achieves significant speedup over existing methods while maintaining clinically acceptable accuracy on diverse real-world imaging. This enables practical deployment in fast-paced clinical environments, speeding up cardiology consultation and facilitating digital ECG storage for retrospective health analysis research.

## Data Availability

All data produced are available online at https://physionet.org/

https://github.com/alphanumericslab/ecg-image-kit

https://physionet.org/content/mimic-iv-ecg/1.0/

https://physionet.org/content/ptb-xl/1.0.3/

## References

[1] Awni Y Hannun, Pranav Rajpurkar, Masoumeh Haghpanahi, Geoffrey H Tison, Codie Bourn, Mintu P Turakhia, and Andrew Y Ng. Cardiologist-level arrhythmia detection and classification in ambulatory electrocardiograms using a deep neural network. Nat. Med., 25(1):65–69, January 2019.

[2] Yanrui Jin, Zhiyuan Li, Mengxiao Wang, Jinlei Liu, Yuanyuan Tian, Yunqing Liu, Xiaoyang Wei, Liqun Zhao, and Chengliang Liu. Cardiologist-level interpretable knowledge-fused deep neural network for automatic arrhythmia diagnosis. Commun. Med. (Lond.), 4(1):31, February 2024.

[3] Konstantinos C Siontis, Peter A Noseworthy, Zachi I Attia, and Paul A Friedman. Artificial intelligence-enhanced electrocardiography in cardiovascular disease management. Nat. Rev. Cardiol., 18(7):465–478, July 2021.

[4] Zachi I Attia, Suraj Kapa, Francisco Lopez-Jimenez, Paul M McKie, Dorothy J Ladewig, Gaurav Satam, Patricia A Pellikka, Maurice Enriquez-Sarano, Peter A Noseworthy, Thomas M Munger, Samuel J Asirvatham, Christopher G Scott, Rickey E Carter, and Paul A Friedman. Screening for cardiac contractile dysfunction using an artificial intelligence-enabled electrocardiogram. Nat. Med., 25(1):70–74, January 2019.

[5] Matplotlib 2014; visualization with python — matplotlib.org. https://matplotlib.org/. [Accessed 09-06-2025].

[6] GitHub - dy1901/ecg_plot: Plot standard multi lead ECG/EKG chart with Python — github.com. https://github.com/dy1901/ecg_plot. [Accessed 09-06-2025].

[7] Kshama Kodthalu Shivashankara, Deepanshi, Afagh Mehri Shervedani, Gari D Clifford, Matthew A Reyna, and Reza Sameni. ECG-Image-Kit: a synthetic image generation toolbox to facilitate deep learning-based electrocardiogram digitization. Physiol. Meas., 45(5):055019, May 2024.

[8] Anthony Demolder, Viera Kresnakova, Michal Hojcka, Vladimir Boza, Andrej Iring, Adam Rafajdus, Simon Rovder, Timotej Palus, Martin Herman, Felix Bauer, Viktor Jurasek, Robert Hatala, Jozef Bartunek, Boris Vavrik, and Robert Herman. High precision ECG digitization using artificial intelligence. J. Electrocardiol., 90:153900, February 2025.

[9] Patrick Wagner, Nils Strodthoff, Ralf-Dieter Bousseljot, Wojciech Samek, and Tobias Schaeffter. PTB-XL, a large publicly available electrocardiography dataset, 2022.

[10] Veer Sangha, Bobak J Mortazavi, Adrian D Haimovich, Antônio H Ribeiro, Cynthia A Brandt, Daniel L Jacoby, Wade L Schulz, Harlan M Krumholz, Antonio Luiz P Ribeiro, and Rohan Khera. Automated multilabel diagnosis on electrocardiographic images and signals. Nat. Commun., 13(1):1583, March 2022.

[11] Antonio Luiz P Ribeiro, Gabriela MM Paixão, Paulo R Gomes, Manoel Horta Ribeiro, Antônio H Ribeiro, Jéssica A Canazart, Derick M Oliveira, Milton P Ferreira, Emilly M Lima, Jermana Lopes de Moraes, Nathalia Castro, Leonardo B Ribeiro, and Peter W Macfarlane. Tele-electrocardiography and bigdata: The CODE (clinical outcomes in digital electrocardiography) study. J. Electrocardiol., 57S:S75– S78, November 2019.

[12] Sagnik Das, Ke Ma, Zhixin Shu, Dimitris Samaras, and Roy Shilkrot. DewarpNet: Single-image document unwarping with stacked 3D and 2D regression networks. In 2019 IEEE/CVF International Conference on Computer Vision (ICCV). IEEE, October 2019.

[13] Felix Hertlein, Alexander Naumann, and Patrick Philipp. Inv3D: a high-resolution 3D invoice dataset for template-guided single-image document unwarping. Int. J. Doc. Anal. Recognit., 26(3):175–186, September 2023.

[14] Vincenzo Randazzo, Edoardo Puleo, Annunziata Paviglianiti, Alberto Vallan, and Eros Pasero. Development and validation of an algorithm for the digitization of ECG paper images. Sensors (Basel), 22(19):7138, September 2022.

[15] Julian D Fortune, Natalie E Coppa, Kazi T Haq, Hetal Patel, and Larisa G Tereshchenko. Digitizing ECG image: A new method and open-source software code. Comput. Methods Programs Biomed., 221(106890):106890, June 2022.

[16] E Morales, D Sevilla,H H Pierluissi, and H Nazeran. Digitization and synchronization method for electrocardiogram printouts. In 2005 IEEE Engineering in Medicine and Biology 27th Annual Conference. IEEE, 2005.

[17] Lakshminarayan Ravichandran, Chris Harless, Amit J Shah, Carson A Wick, James H Mcclellan, and Srini Tridandapani. Novel tool for complete digitization of paper electrocardiography data. IEEE J. Transl. Eng. Health Med., 1:1800107–1800107, 2013.

[18] Siddharth Mishra, Gaurav Khatwani, Rupali Patil, Darshan Sapariya, Vruddhi Shah, Darsh Parmar, Sharath Dinesh, Prathamesh Daphal, and Ninad Mehendale. ECG paper record digitization and diagnosis using deep learning. J. Med. Biol. Eng., 41(4):422–432, June 2021.

[19] Yao Li, Qixun Qu, Meng Wang, Liheng Yu, Jun Wang, Linghao Shen, and Kunlun He. Deep learning for digitizing highly noisy paper-based ECG records. Comput. Biol. Med., 127(104077):104077, December 2020.

[20] Brian Gow, Tom Pollard, Larry A Nathanson, Alistair Johnson, Benjamin Moody, Chrystinne Fernandes, Nathaniel Greenbaum, Jonathan W Waks, Parastou Eslami, Tanner Carbonati, Ashish Chaudhari, Elizabeth Herbst, Dana Moukheiber, Seth Berkowitz, Roger Mark, and Steven Horng. MIMIC-IV-ECG: Diagnostic electrocardiogram matched subset, 2023.

[21] Blender Foundation. blender.org - Home of the Blender project - Free and Open 3D Creation Software — blender.org. https://www.blender.org/. [Accessed 09-06-2025].

[22] Dustin Podell, Zion English, Kyle Lacey, Andreas Blattmann, Tim Dockhorn, Jonas Müller, Joe Penna, and Robin Rombach. SDXL: Improving latent diffusion models for high-resolution image synthesis. 2023.

[23] Hu Ye, Jun Zhang, Sibo Liu, Xiao Han, and Wei Yang. IP-Adapter: Text compatible image prompt adapter for Text-to-Image diffusion models. 2023.

[24] Roman Suvorov, Elizaveta Logacheva, Anton Mashikhin, Anastasia Remizova, Arsenii Ashukha, Aleksei Silvestrov, Naejin Kong, Harshith Goka, Kiwoong Park, and Victor Lempitsky. Resolution-robust large mask inpainting with fourier convolutions. In 2022 IEEE/CVF Winter Conference on Applications of Computer Vision (WACV). IEEE, January 2022.

[25] Gao Huang, Zhuang Liu, Laurens van der Maaten, and Kilian Q Weinberger. Densely connected convolutional networks. In 2017 IEEE Conference on Computer Vision and Pattern Recognition (CVPR). IEEE, July 2017.

[26] Ultralytics. YOLO11 NEW — docs.ultralytics.com. https://docs.ultralytics.com/models/yolo11/. [Accessed 10-06-2025].

[27] Andrej Iring, Viera Krešňáková, Michal Hojcka, Vladimir Boza, Adam Rafajdus, and Boris Vavrik. PMcardio ECG image database (PM-ECG-ID): A diverse ECG database for evaluating digitization solutions, 2024.

[28] Jian Song, Hongruixuan Chen, and Naoto Yokoya. Syntheworld: A large-scale synthetic dataset for land cover mapping and building change detection. In Proceedings of the IEEE/CVF Winter Conference on Applications of Computer Vision, pages 8287–8296, 2024.

[29] Tin Barisin, Christian Jung, Anna Nowacka, Claudia Redenbach, and Katja Schladitz. Cracks in concrete. arXiv [cs.CV], 2025.

[30] Rodrigo Rill-García, Eva Dokladalova, and Petr Dokládal. Syncrack: Improving pavement and concrete crack detection through synthetic data generation. In Proceedings of the 17th International Joint Conference on Computer Vision, Imaging and Computer Graphics Theory and Applications. SCITEPRESS — Science and Technology Publications, 2022.

[31] Klaus Greff, Francois Belletti, Lucas Beyer, Carl Doersch, Yilun Du, Daniel Duckworth, David J Fleet, Dan Gnanapragasam, Florian Golemo, Charles Herrmann, Thomas Kipf, Abhijit Kundu, Dmitry Lagun, Issam Laradji, Hsueh-Ti Liu, Henning Meyer, Yishu Miao, Derek Nowrouzezahrai, Cengiz Oztireli, Etienne Pot, Noha Radwan, Daniel Rebain, Sara Sabour, Mehdi S M Sajjadi, Matan Sela, Vincent Sitzmann, Austin Stone, Deqing Sun, Suhani Vora, Ziyu Wang, Tianhao Wu, Kwang Moo Yi, Fangcheng Zhong, and Andrea Tagliasacchi. Kubric: A scalable dataset generator. In 2022 IEEE/CVF Conference on Computer Vision and Pattern Recognition (CVPR). IEEE, June 2022.

[32] Carl Doersch, Ankush Gupta, Larisa Markeeva, Adria Recasens, Lucas Smaira, Yusuf Aytar, Joao Carreira, Andrew Zisserman, and Yi Yang. TAP-vid: A benchmark for tracking any point in a video. Advances in Neural Information Processing Systems, 35:13610–13626, 2022.

[33] Felipe Rodrigues Perche Mahlow, André Felipe Zanella, William Alberto Cruz-Castañeda, and Marcellus Amadeus. An inpainting-infused pipeline for attire and background replacement. ArXiv, abs/2402.03501, 2024.

